# User experiences with a digital health family planning counseling tool for women living with HIV in Kenya

**DOI:** 10.64898/2025.12.22.25342549

**Authors:** Agnes Karingo Karume, Alison L. Drake, June Moraa, Celestine Atieno, Nancy Ngumbau, Aparna Seth, Kristin Beima-Sofie, John Kinuthia, Jennifer A. Unger

## Abstract

Women with HIV(WLHIV) have diverse and complex reproductive health needs that require patient-centered, informed decision making. Digital tools to support reproductive life planning may improve reproductive health counseling and outcomes for these women. We evaluated user-experiences with a self-administered, tablet-based family planning(FP) counseling tool as part of a packaged digital health intervention evaluated in a cluster randomized trial in Kenya.

We conducted focus group discussions with WLWH who received the counseling tool(n=10) and in-depth interviews with providers(n=10). Data were collected using semi-structured guides, transcribed, translated, and thematically analyzed.

Among 95 women, median age was 26 years and 52% were married. Providers had median 7 years’ experience. WLWH and providers found the counseling tool acceptable, and felt it improved decision-making, FP knowledge, and patient-provider interactions. Women reported the counseling tool supported informed FP decisions, educated on less familiar options, and dispelled myths and misconceptions. Providers said the counseling tool helped tailor counseling though it was challenging to use with women with more FP experience or with a selected method, but helpful among FP-initiators and adolescents.

The counseling tool was useful and acceptable, but too lengthy. Tailoring the counseling tool and making it optional for specific groups of WLWH may improve feasibility.

## Background

Meeting the family planning (FP) needs of women living with HIV (WLHIV) is essential for advancing reproductive rights, improving maternal and child health, and preventing vertical transmission of HIV^1^. Unmet need for FP among WLHIV persists in sub-Saharan Africa, with rates around 30%^2,^ and unintended pregnancy rates as high as 40%^3,4^. Many WLHIV continue to experience substantial gaps in access to modern contraception due to stigma, limited method choice, fragmented service delivery, and inadequate integration of FP within HIV care settings. WLWH also confront misinformation, fear of side effects, and limited knowledge about contraceptive options^5,6^. Strengthening contraceptive counseling, particularly through evidence-based and client-centered approaches, may help reduce unmet need for FP, support reproductive autonomy, and ultimately prevent adverse outcomes related to unintended pregnancy^6^.

Routine HIV care clinics offer a strategic opportunity to address the contraceptive needs of WLHIV. Although more than 70% of WLHIV in sub-Saharan Africa wish to delay or avoid future pregnancies, many do not receive adequate FP support within their routine HIV care^6,1^. Contraceptive use in this population remains suboptimal, with persistent reliance on short-acting, user-dependent methods such as injectables and oral pills, which are more susceptible to discontinuation and inconsistent use^,8^. Long-acting, reversible contraception (LARC) are highly effective and user-independent and may be suitable for women who desire longer term, reliable protection against pregnancy. However, LARC are underutilized in African settings due to provider bias, misinformation, and limited availability within HIV clinics^7,8^Importantly effectiveness alone may not drive women’s contraceptive choices. It is critical to ensure that women are supported to make contraceptive choices that reflect their own values, preferences, and reproductive goals. This includes access and support to use the full range of methods, short and long-acting, while addressing barriers that constrain choice. Closing these gaps through integrated, rights-based contraceptive counseling within HIV care could enhance informed method choice and support WLHIV in achieving their reproductive goals.

Digital health innovations are increasingly being used to strengthen FP counseling in low- and middle-income countries. More recent efforts have focused on interactive digital decision support and counseling tools designed to support informed contraceptive choice^9,10^. Tablet and computer-based decision aids have been shown to improve women’s knowledge, dispel myths and support shared decision making in diverse settings^9,11,10^. In high income countries, tools such as Smart Choices^12^, My Birth Control^13^, MyPath^14^ and Tuune^11^ have demonstrated that digital counseling interventions can improve method knowledge, support selection of methods consistent with preferences and increase satisfaction with contraceptive counseling^15,10^. However, rigorous evaluations of client-facing digital health tools that support informed contraceptive decision-making in LMICs, and particularly for WLHIV, remain limited. Additionally, most platforms have primarily emphasized medical eligibility criteria (MEC) rather than incorporating women’s individual values and preferences into method selection. Examples include programmatic roll-outs of the WHO-MEC application ‘MEC wheel’^16^ and digital job-aids derived from the Balanced Counseling Strategy^17,18^ which support rapid screening and method safety but do not account for contextual factors such as fertility intentions. Digital counseling tools that explicitly integrate women’s preferences alongside clinical eligibility may better match WLHIV to methods they can continue using, potentially reducing discontinuation and unmet need.

Little is known about how WLHIV specifically experience digital FP counseling tools or whether digital FP counseling interventions address their unique reproductive health priorities. Although several digital tools have been developed to support contraceptive counseling^10,15^, few have been rigorously evaluated for their effectiveness among WLHIV, particularly within routine HIV care settings. WLHIV often encounter barriers such as stigma, inconsistent counseling, and limited privacy, which can hinder informed contraceptive decision-making. In Kenya, most routine HIV care is documented in point-of-care electronic medical records, primarily the KenyaEMR^19,20^. Within this system, the provider form captures FP information, with FP status designated as a mandatory field, reinforcing documentation during ART visits. However, the information captured is minimal and limited to FP status and method choice, which does not reflect comprehensive counseling, fertility intentions, or client preferences, leaving significant gaps in addressing the reproductive health needs of WLHIV.

Self-administered, client-facing tools that provide individualized information and allow users to engage at their own pace may help overcome these barriers and complement provider-delivered counseling. In response to this need, we developed a tablet based counseling tool that places women’s preferences at the center of contraceptive decision-making, alongside clinical considerations to support informed, values based FP decision making. Understanding user experiences with the tool is essential to refine counseling approaches that promote informed choice and support reproductive autonomy among WLHIV. To this end, we evaluated user-experiences with the patient-facing, self-administered tool as part of a digital intervention being evaluated in a cluster randomized trial in Kenya^21^.

## Methods

### Study design and setting

Between June and July 2024, we conducted in-depth interviews (IDIs) with HIV care providers offering RH/FP services (n=10) and ten focus group discussions (FGDs, 8-10 WLWH/FGD) with a subset of WLWH participating in cluster randomized clinical trial of a digital health intervention, Mobile WACh Empower. Participants were recruited from intervention sites located at five Kenyan HIV and FP clinics: two rural/ peri-urban sites in Western Kenya and three periurban/urban sites in Nairobi. All facilities were public and served over 1000 people living with HIV.

### RH decision-support counseling tool

The Mobile WACh Empower trial is evaluating a combined digital health intervention: a self-administered, client-facing, tablet-based RH decision-support counseling tool (CT), self-administered at baseline coupled with follow-up, tailored text messages delivered over two years^21^. WLWH completed the CT independently during routine clinic visits, prior to seeing a clinician or nurse and the trial study visit, with study staff available for assistance if needed. The CT includes medical eligibility criteria for contraceptive use (including compatibility with antiretroviral therapy), fertility intentions, prior experience with contraception, as well as values and preferences for reproductive life planning including (but not exclusive to) contraception. Counseling content is subsequently tailored based on individual responses using a programmed algorithm. The tool provides tailored information on specific FP methods and includes educational content that addresses myths and misconceptions related to FP (Fig 1). The CT assesses desire for pregnancy and captures timing of desired pregnancy. If women are unsure about using contraception but are not currently planning a pregnancy, the CT offers information on short-acting or user-controlled methods that can be discontinued at any time.

**Figure 1.**
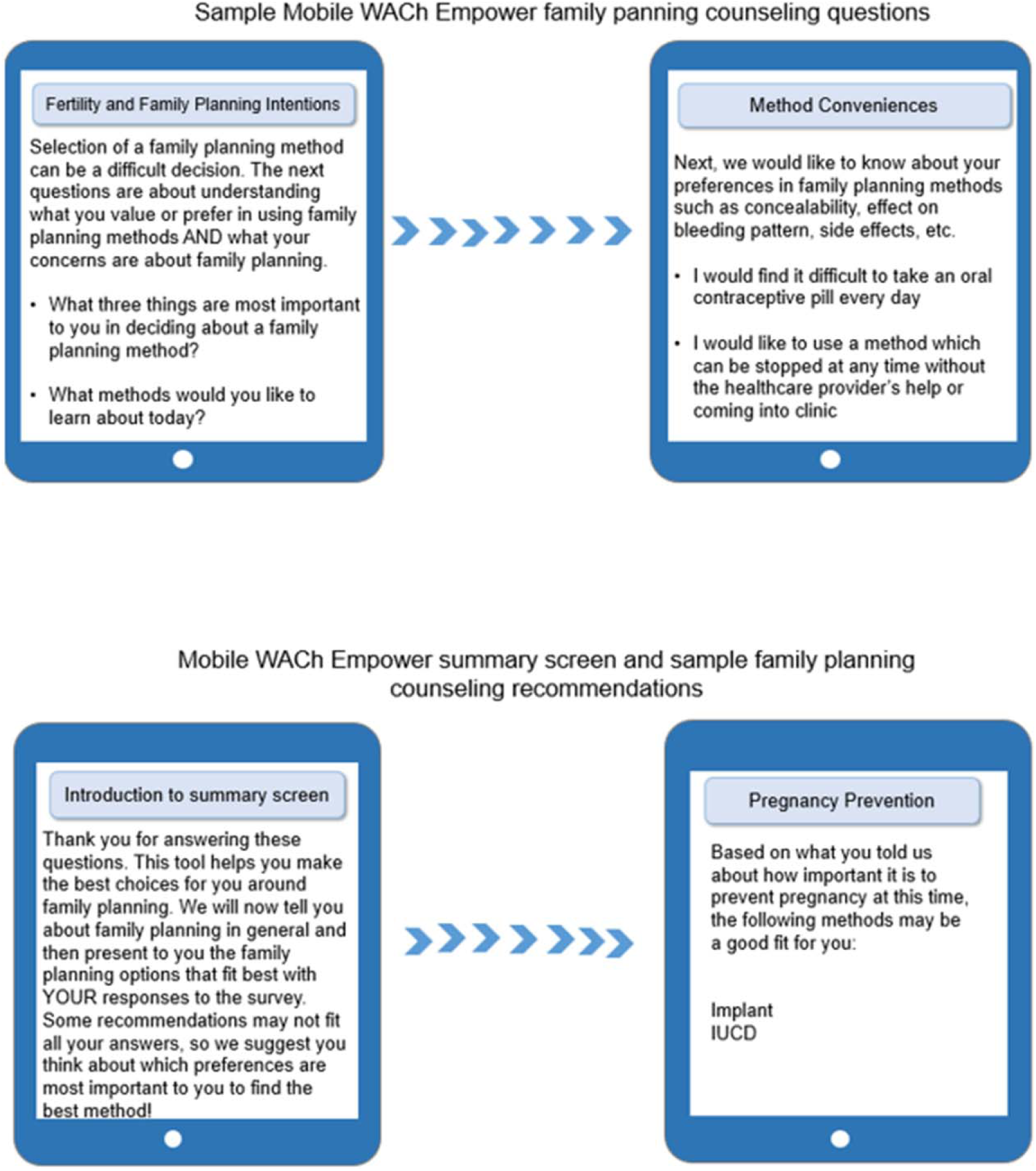
Mobile Wach Empower Family Planning Counseling Tool.

At the end of the CT, an automated summary screen was generated based on the participants’ prior responses. For women who desire pregnancy prevention, the CT summary screen concludes with recommendations for FP method(s) that are appropriate based on participant’s responses and stratified by the criteria under which the method is suitable, such preferences for dosing, effectiveness, or duration of coverage (Fig 2). For women who desire pregnancy, the CT summary screen presents information on safe conception and pregnancy, including the importance of maintaining a low HIV viral load, continuing ART, safety of dolutegravir, early antenatal care, exclusive breastfeeding, and optimal birth spacing. The CT summary screen was printed by a study staff member, which was given to the HIV care provider prior to their routine care visit and RH/FP counseling.

**Figure 2.**
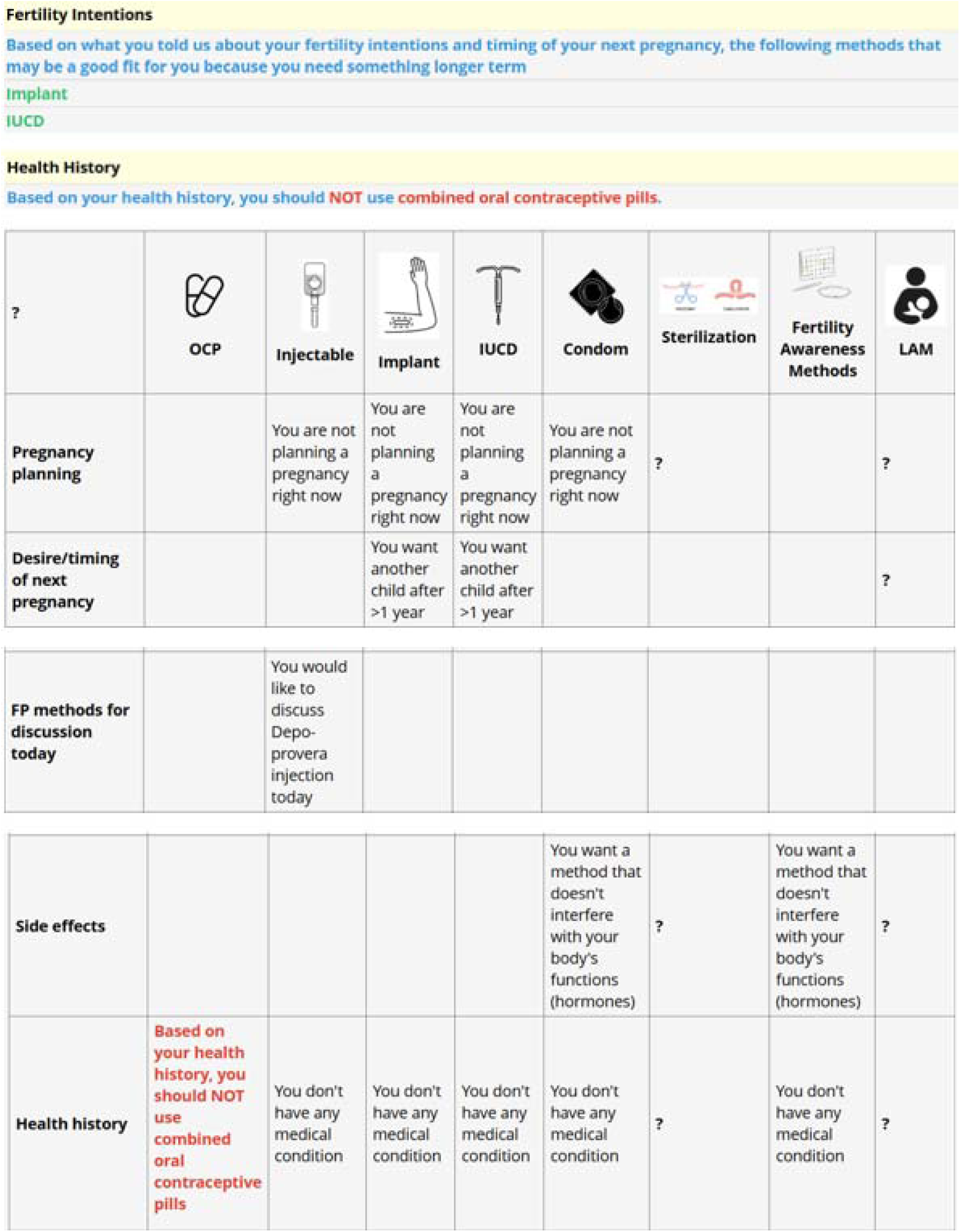
Mobile Wach Empower Family Planning Summary Screen.

Women were eligible for trial participation if they were WLHIV of reproductive age (18–45; 14–17 years if emancipated minor), receiving HIV care at study site and planning to receive care at the enrollment facility for 2 years, had daily access to a mobile phone (own or shared) with Safaricom SIM, and were literate or comfortable with someone reading the study SMS. Pregnant women were ineligible.

### Study recruitment and procedures

After trial enrollment was completed, study staff recontacted women enrolled in the trial at intervention sites who had previously interacted with the CT; recontact occurred within 15 months of enrollment. Women were purposively sampled, stratified by age (≤25 and >25 years) and FP experience/ fertility intentions. We initially contacted women who had engaged with the counselling tool within the past six months; however, we were unable to recruit sufficient numbers for the FGDs and subsequently broadened the eligibility criteria to include participants who had engaged with the CT at any point following enrollment; all participants were enrolled in the trial within the past 15 months. Providers were eligible for participation if they provided HIV care at intervention sites and had been asked to use the CT summary screen; they were also recruited after completion of trial enrollment. Women and providers interested and eligible provided written informed consent prior to enrollment.

During FGDs, women were refamiliarized with the CT by being shown the paper version and given time to walk through the CT questions before providing feedback on their experiences using it. In IDIs, providers offered feedback on their perceptions of women’s use of the CT and their own experiences using a summary screen (generated after completing the CT) during counseling sessions. While the FGD and IDI discussion guides offered the opportunity for participants to provide feedback on content and the CT summary screens on both contraception and planning for pregnancy; participants did not directly comment on pregnancy planning. Therefore, results primarily focus on feedback related to contraceptive counseling.

Additionally, providers completed a post-intervention survey assessing acceptability, appropriateness and feasibility of the CT using the Acceptability of Intervention Measure (AIM), Intervention Appropriateness Measure (IAM), and Feasibility of Intervention Measure (FIM)^22^. Each measure is a 4-item scale with a score ranging from 1-4. All IDIs and FGDs were conducted by trained Kenyan qualitative researchers, audio-recorded, transcribed into English, Dholuo, or Swahili, and translated into English by FGD facilitators.

### Data analysis

We conducted an inductive thematic analysis. Three coders (CA, JM, AK) developed a codebook with eight code groups and 58 descriptions. Each coder independently conducted primary and secondary coding of transcripts. Discrepancies were resolved through discussion and consensus, with input from a senior qualitative researcher (KBS) as needed. Following coding, we generated queries to bring together data coded similarly across transcripts. These queries were summarized into theme-based memos, which helped the research team identify and refine major themes and sub-themes. We used these memos to compare, align, and organize findings, which formed the basis of the final results. All analysis was managed using ATLAS.ti version 24.

### Ethical considerations

This study was approved by the Kenyatta National Hospital –University of Nairobi Ethics Committee (P162/03/2022) in Kenya and the University of Washington Institutional Review Board (STUDY00013136).

## Results

A total of 95 WLHIV participated in FGDs, with median age of 26 years (interquartile range [IQR]: 24–34). Half (53%, n=50) were married, and 58% (n=55) had secondary education or less (Table1). Overall, 78% (n=74) used FP, including 6% (n=6) who were newly initiating a method. Over half of women (55%, n=52) were not planning a pregnancy, 39% (n=37) stated desire for a future pregnancy, and 6% (6) held ambivalent feelings about a future pregnancy (Table 1).

**Table 1.**
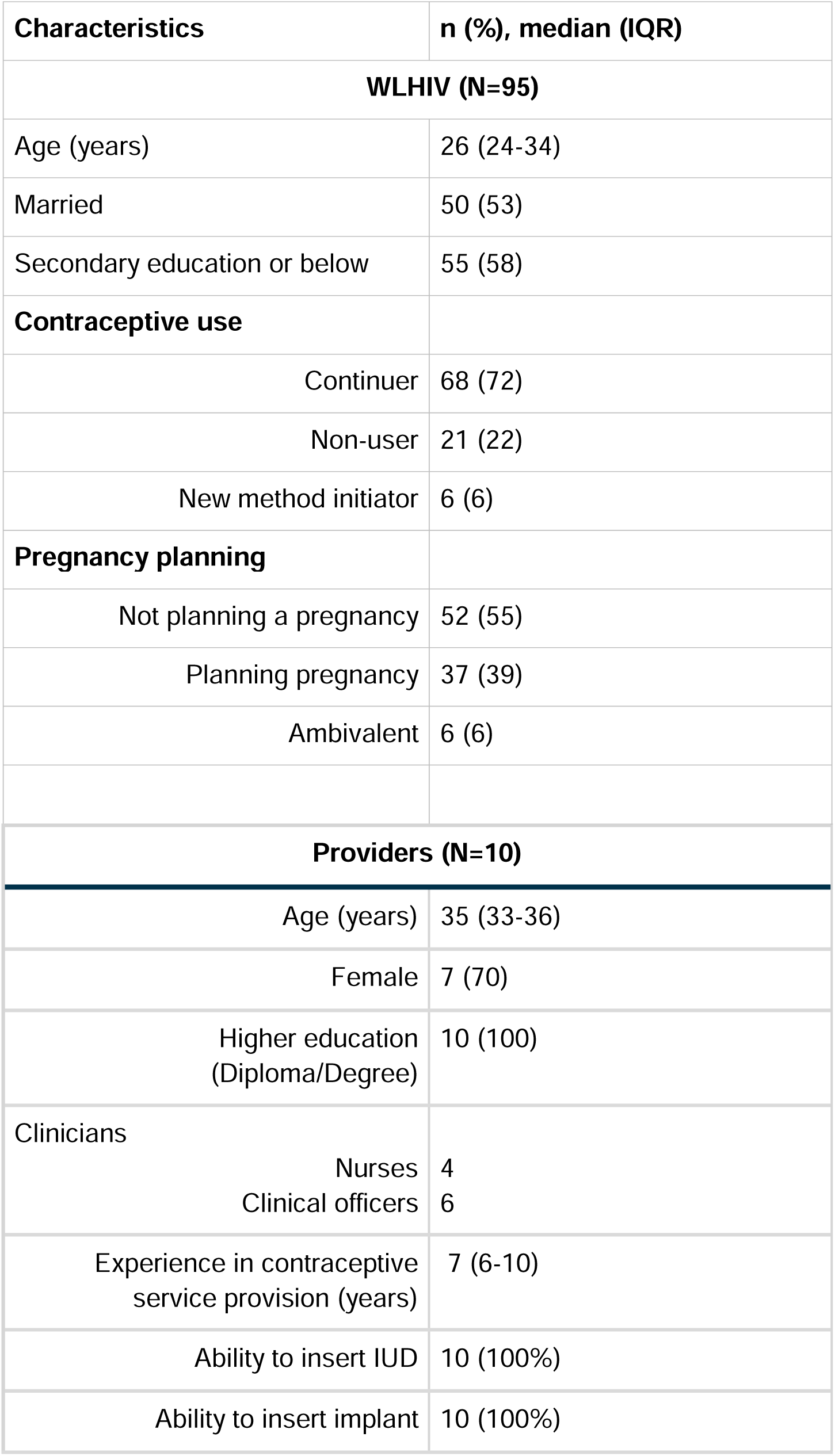
Descriptive characteristics of women living with HIV (WLHIV) (*N*_=_95) and health care providers (*N*_=_10)

Among 10 providers who participated in IDIs, the median age was 35 years (IQR: 33–36), and 70% (n=7) were female (Table 2). All providers had at least a diploma or a degree, with a median of 7 years (IQR: 6–10) of experience in contraceptive service provision. All reported being able to insert both intrauterine devices (IUDs) and implants.

**Table 2.** Women Living with HIV’s Reported Acceptability, Appropriateness and Feasibility of Counseling Tool.

| **Acceptability (AIM)** | **Mean** | **SD** |
| --- | --- | --- |
| Meets approval | 4.70 | 0.48 |
| Appealing | 4.30 | 0.67 |
| Likable | 4.50 | 0.53 |
| Welcome | 4.70 | 0.48 |
| Overall- AIM | 4.55 | 0.35 |
| **Appropriateness** | **Mean** | **SD** |
| Fitting | 4.10 | 0.32 |
| Suitable | 4.10 | 0.57 |
| Applicable | 4.10 | 0.74 |
| Good match | 4.10 | 0.74 |
| Overall- IAM | 4.10 | 0.52 |
| **Feasibility** | **Mean** | **SD** |
| Implementable | 4.20 | 0.63 |
| Possible | 4.10 | 0.57 |
| Doable | 4.10 | 0.57 |
| Easy to use | 4.20 | 0.63 |
| Overall-FIM | 4.15 | 0.57 |

### Perceived acceptability and usefulness of the counseling tool on contraception

Women generally found the CT acceptable and comprehensive, and thought it provided extensive information on FP methods. They found the CT was helpful in providing information on potential side effects of FP methods to make informed decisions. Many women acknowledged they had previously been apprehensive about contraceptive side effects, such as weight gain, menstrual irregularities, and changes in libido. However, women said the CT helped them understand which side effects were temporary, manageable, and/or less common.

*“This tool has helped me to know methods of family planning that I can select from. It also helped me to know how to plan well for my next baby. It is not just a matter of giving birth without a plan. This tool is so helpful … There is the knowledge you are getting from it.” _ WLHIV*

Providers agreed that the counseling tool was comprehensive and provided information, offering preliminary information about options for FP and identifying needs for further counseling, including counseling on side effects and methods that may be well suited to women’s individual needs, preferences, and medical eligibility.

“*I would strongly agree that the counseling needs of the clients are well met… they are able to relate, get in touch with different family planning methods, their availability, side effects, and compatibility*.” _ Provider with 11 years of experience providing FP services

Women also felt the CT addressed common myths, such as FP delaying fertility, causing infertility, or IUCDs relocating elsewhere in the body, and facilitated conversations to dispel additional myths with providers.

“*I was afraid that if I used the injection, I would never conceive again, but the provider used the tool to show me that it’s reversible”_ WLHIV*

Providers agreed that the CT seemed to help dispel myths about FP prior to their counseling sessions. Providers appreciated that they did not need to discuss every FP method in detail, as the CT had already guided clients through the options. Providers felt this allowed consultations to be more focused on addressing specific questions and finalizing method selection.

*“Many women came in fearing that family planning causes cancer or permanent infertility. With the tool, we could break down the science and reassure them with facts”*.

*_* Provider with 11 years of experience providing FP services

Women said the CT was useful in preparing them with relevant information before engaging with providers. They also reported feeling reassured and were more confident in deciding to initiate or continue their selected method of FP after using the CT. Women perceived the CT facilitated their decision making in choosing an appropriate method or switching to a method better suited to them. Women who were currently using FP said the information from the CT helped them learn about other methods they were not familiar with and, for some, facilitated switching to another method that might be more suitable for them. One woman explained,

*“All along I knew that Depo is the only family planning method, and I did not know of any other method. When I came here, I was given the tool and I looked at it and read through it. I read everything when I learned that I would not use Depo anymore and I was going for an implant because I observed that the implant was being inserted in the upper arm… I was psychologically prepared for the method that the nurse would give me how it looked, and where it would be inserted”. _WLWH*

While providers did not use the CT with women, they were trained on it and were familiar with the content. Providers perceived the CT to flow well, with logical, algorithm-based sequential questions that acted as guide for women, tailoring counseling content and preparing women to make informed decisions. Providers thought women appreciated these features. *“*

*“The tool acts as a guide, spreading out throughout the different locally available methods of family planning to create awareness for the client and to prepare them in terms of making choices.” _*Provider with 13 years of experience providing FP services

Providers also noted that the CT facilitated patient-provider interactions by fostering closer relationships and enhancing rapport.

*“This tool helps you get closer to your client. It brings you closer. It even increases the rapport between clients and providers because you spend more time together, and it gives the client confidence in you as a healthcare worker.” _* Provider with 5 years of experience providing FP services

Several women liked the ability to self-administer the CT, noting that it allowed them to respond to questions privately and comfortably, without feeling embarrassed. Providers also noted the potential for privacy afforded by the CT being self-administrated.

Providers felt the CT helped women talk with partners about FP, supporting the belief that FP was “okay” to use. However, women had varied perceptions of the role of the CT in involving male partners. Some women said the CT played a role in making informed decisions with their partners, since they were able to relay information from the CT to their partner, enabling them to decide on a method as a couple. However, other women found that the information from the CT helped them select a discreet FP method, circumventing partner concerns about FP:

“*I … had problems with my partner…and the provider helped me and even told me that if that is the case then you can use something that is discreet.” _WLHIV*

However, younger women had alternative views; they did not think partner-related factors should be included in the CT since they felt FP was their decision, and partners did not need to be involved:

*“I am meeting with him today, I will even use a condom, that is none of his business. We should not be asked [in the CT] about partner status because even himself, my family planning choice is none of his business.” _WLHIV*

Women found the CT was an acceptable mechanism to deliver counseling and suggested that it could be beneficial to a wider range of women, not just WLWH. They noted it would, however, require changes to the language to make the CT more inclusive, such as removing HIV-specific content.

*“So, it can be adapted if it can be generalized. This section about HIV is to be removed. Then it will be adopted by the public or general population”. _WLWH*

### Perceived acceptability and usefulness of the summary screen on contraceptive decision-making

Women said the CT summary screen prepared them to ask questions about their concerns for counseling with their providers which facilitated FP counseling more aligned to their needs.

Women said they felt more informed and aware about FP methods and side effects, which made them more comfortable and better prepared to engage with providers.

*“Concerning using the summary screen, it prepared me to ask if the method I was using that was disturbing me, if I can continue with it and if it will work.” _WLHIV*

Women also appreciated the visual depictions of the FP methods on the CT summary screen and suggested adding additional visuals to the other components of the CT to support method comprehension.

*“…your mind will be able to understand what it can see. Maybe I don’t even know what an implant is, so even starting to read about it and you don’t even know it, is hard, but if you draw it, maybe I even know it, but it is the name that I don’t know.” _WLHIV*

Providers described the CT summary screen as acceptable, simple, easy to use, and said that it mirrored activities they were tasked with providing. Providers felt that the CT summary screen streamlined their workflow, as some women who had used the CT had already made informed decisions about their preferred FP method. They thought the summary information in the CT summary screen eliminated the need to review all FP methods, allowing them to focus on confirming method choice and addressing any specific concerns.

*“The client comes in with a clear decision—she has already chosen the method she wants to use. This makes my work easier, as I don’t have to go through the entire family planning process with her.” _*Provider with 5 years of experience providing FP services

While some providers said the CT did not significantly impact their workflow, others felt workload was higher if the CT summary screen directed providers to revisit and discuss more FP methods than they might have otherwise if women required additional guidance based on their preferences or medical eligibility.

Providers said they used the CT summary screen to assess fertility intentions and discuss FP method eligibility; these topics were acceptable to inquire about as they were already captured in the electronic medical record (EMR) platform used to deliver routine services. Providers noted that the CT covered all modern methods, and they used the CT summary screen to provide information to women about the different methods that women enquired about or expressed interest in. If women were currently using FP, providers said they focused counseling on these methods instead of the other methods on the CT summary screen. However, providers did acknowledge that they did not use the CT summary screen with all women; they used it primarily with women who were contraceptive-naive or adolescents, as both groups were perceived to have limited knowledge about FP. Providers also felt compelled to provide comprehensive counseling to adolescents to help curb high adolescent fertility rates. Providers said the amount of time they spent using the CT summary screen varied based on women’s prior experience with FP, with more time spent with women who were contraceptive-naive.

*“For a naïve patient, it’s prudent that you just sit down and take your time and check them through the whole content. But if the patient is already familiar, you just remind them about the options available…It also differs from client to client, there are those clients who maybe are naïve about the family planning methods, there are those who have information that really fits, so you don’t go into so much elaboration.” _*Provider with 11 years of experience providing FP services

Providers did not perceive the CT summary screen to be useful for women who were post-menopausal, or using permanent or long-term FP methods. They also said it was challenging to use with women who either had significant experience using a specific FP method or had already identified a preferred method and were satisfied with their selection; and opted not to use the CT summary screen with these women.

### Perceived implementation considerations and recommendations

Providers reported high acceptability (mean=4.55, standard deviation [SD]=0.35), appropriateness (mean=4.10, SD=0.52), feasibility (mean=4.15, SD=0.57)(Table 3). However, despite support from women and providers to use the CT and the summary screen to guide counseling, several implementation considerations were mentioned that may influence consistent and effective use.

Providers noted differences between the CT and the existing Kenyan EMR, which only assesses three components of reproductive health: pregnancy status, desire for pregnancy, and method of FP used (if any). Providers thought the CT could serve as an expansion of the EMR and mentioned the ability to assess side effects as a key advantage of the platform.

*“The EMR prompts us to ask about pregnancy intention and method use, but this tool goes further—it helps us explore side effects, medical eligibility, and client concerns more systematically.“_* Provider with 5 years of experience providing FP services

However, based on the providers’ familiarity with the CT, they also felt the CT algorithm could be more tailored to better individualize and direct women to specific information needed. As the EMR does not prompt providers to capture data or guide providers to ask specific questions, this functionality was perceived to be desirable by providers. Providers thought that integrating the CT into existing systems, such as the EMR, would be helpful as it would avoid the need to switch platforms while delivering care and services, and prevent redundancies between platforms. Providers were supportive of integrating the CT into routine HIV care provided by the Ministry of Health.

While women appreciated the comprehensiveness of the CT, they also unanimously stated it was too long, and some questions were reported to feel repetitive and time consuming. For less literate women, the CT took longer to administer either because women needed more time to read and digest the content, or because they required additional assistance from a study staff member to use it.

“*The tool was too long, and then it had some challenging questions because you can find there is a question you were asked earlier, and when you move forward, you find another question that looks like the one you already answered*.“_ *WLHIV*

Women suggested modifying the CT by formatting content as questions rather than statements to promote better comprehension. While providers said the language in the CT was not very technical, they also acknowledged that the CT would be challenging for a low literacy population, and that these women may need assistance with a provider as they would not be able to self-administer the CT. Additional challenges with the network were also cited, as the CT initially required consistent internet access, which meant that the CT would need to be restarted if internet connectivity was lost.

Time to counsel women using the CT summary screen was also reported to be variable; providers reported spending 30-60 minutes counseling women not familiar with FP, but significantly less time (5-10 minutes) with women familiar with FP. While some providers felt the CT reduced the amount of time required to counsel on methods, other providers thought it could increase time if women wanted to discuss many methods from the summary screen or many concerns. Sometimes, providers acknowledged not using the CT summary screen to focus on delivering essential services that were perceived to be higher priority, particularly when facilities were short staffed. If there were long patient queues in the clinic, both women and providers concurred that the CT summary screen was less likely to be used. Providers said this was especially true if there were multiple FP methods on the CT summary screen. Some providers modified how they used the CT, and subsequently counseled women, based on patient volume and time constraints, prioritizing key aspects of the summary screen rather than going through the entire summary. They recommended increasing sensitization among clinicians and staff at various service points to enhance the tool’s utilization.

*“There are days when we have too many clients, and using the tool in its entirety may not be feasible. In such cases, I focus on the essential components, like fertility intentions and method compatibility, to ensure the client still receives comprehensive care.“* _ Provider with 10 years’ experience in providing FP services

Women expressed discomfort when the CT asked questions about their sexual history and activity, citing concerns related to privacy and fear of judgment. These questions assessed FP use (including condom use), risk perceptions for HIV/sexually transmitted infections, and partner communication. However, they recommended adding content related to mental health in the CT and thought the CT summary screen should include mental health to prompt providers to inquire about this topic.

Women stated that providers may not use the CT summary screen to counsel women on topics they were interested in discussing, such as specific side effects, or they may opt-out of using it entirely. Providers mentioned that the CT does not address stockouts or whether available providers are trained to administer methods; instead, it provides information about FP methods, permitting women to choose whether they want to select another method or go to another facility to obtain the method. Finally, since the CT does not assess all MEC, providers mentioned that they would still be tasked with completing MEC assessments, and may need to use other resources, like the WHO MEC wheel, in conjunction with the CT.

## Discussion

This study highlights how a self-administered, tablet-based counseling tool can enhance FP conversations for WLWH in Kenya. Both women and providers described the tool as acceptable, useful, and empowering. Women noted it gave them new knowledge, dispelled fears, and prepared them to make confident, informed decisions. Providers appreciated that it helped clients arrive at consultations better prepared, focusing discussions on specific concerns rather than repeating information. At the same time, both groups pointed to important challenges: the tool was often too long, difficult for women with lower literacy, and hard to fit into busy clinics where providers are balancing HIV care and reproductive health responsibilities.

Our findings reinforce evidence from digital counseling tools in other settings, which consistently show improvements in women’s knowledge and satisfaction with FP services^9,11,10^. Tools such as My Birth Control^13^ and MyPath^23^ have demonstrated that interactive, client-facing approaches can dispel myths and build confidence in contraceptive choices. What sets our tool apart is its explicit focus on WLWH, addressing their unique clinical considerations, such as ART compatibility, while also centering on women’s values and preferences. By combining medical eligibility with women’s lived realities, the tool offered more individualized guidance than many existing platforms, and in doing so, may help women see themselves reflected in their contraceptive choices.

Notably, women described how the tool created space for them to revisit long-held assumptions. Some women who had relied on injectables for years discovered methods they had never considered, while others realized side effects they once feared were either temporary or uncommon. For adolescents and FP-naïve women in particular, the tool seemed to act as an entry point into a conversation many had never had before. In contrast, women who were already comfortable with their method, or who had long-term or permanent contraception, found the tool less relevant. This distinction underscores a critical point: one size does not fit all.

Global guidance and reviews emphasize tailored, person-centered FP counseling rather than uniform delivery, especially for adolescents^24,25,26^. A streamlined or modular version, with opt-out pathways for experienced users, could make counseling more efficient without diluting its benefits for those who need it most.

Providers also pointed to the dual impact of the summary screen. It often saved time, since clients came in with clearer preferences and questions. However, it sometimes lengthened visits when women wanted to explore multiple options in detail. These experiences suggest that the tool’s value is not just in providing information, but in reframing how women and providers engage with one another shifting consultations from directive to collaborative. Integrating the tool into existing electronic medical records such as the KenyaEMR^20^ which already captures FP status could further streamline workflows and ensure that counseling content is aligned with routine HIV care, minimizing duplication and making it easier to sustain use in the long term.

This is consistent with WHO guidance to integrate digital tools into interoperable health information systems and efforts for HIV and family planning integration^27,28,29^.

Finally, the counseling tool supported women’s autonomy. Many participants described feeling empowered to make voluntary, informed decisions, sometimes in partnership with their partners, and at other times deliberately independent from them. These varied perspectives reflect the realities of WLWH navigating complex social and relational dynamics. Digital tools that allow women to engage privately, at their own pace, may be particularly powerful in giving women control over reproductive decisions, while still leaving room for couple-based counseling when desired^30,13^.

This study has several strengths, including diverse perspectives from both clients and providers, the use of validated measures of acceptability, appropriateness, and feasibility, and the ability to situate findings within the context of an ongoing trial. Limitations include limited discussion of pregnancy planning content, potential recall or social desirability bias, and the fact that participants were drawn from research settings, which may not fully mirror routine practice.

Looking ahead, future work should evaluate whether the tool improves contraceptive uptake, continuation, and satisfaction, and should explore strategies for adapting content to different groups of women. Integration into EMRs and routine HIV care will be essential for scale-up, as will adaptations for adolescents, women not living with HIV, and populations with lower literacy. Equally important is balancing comprehensiveness with efficiency: tailoring content to women’s needs could ensure that the tool is both impactful and sustainable in overstretched clinic environments.

In conclusion, this counseling tool shows promise in transforming FP counseling for WLWH. By equipping women with knowledge, confidence, and voice, it not only improved individual decision-making but also shifted the quality of interactions between women and providers. With refinements to streamline use and better target those who benefit most, the tool has the potential to enhance reproductive autonomy while strengthening integration of HIV and FP services.

## Author contributions

Conceptualization: ALD, JAU, JK Formal analysis: AKK, JM, CA Methodology: ALD, KBS Supervision and validation: ALD, JK, KBS Writing original draft: AKK Writing- review & editing: AKK, ALD, JM, CA, NN, AS, KBS, JK, JAU

## Data Availability

All data produced in the present study are available upon reasonable request to the authors

